# Feasibility, Effectiveness, and Cost-utility of Implementing a Reconsolidation-Based Trauma Treatment in the Aftermath of the November 13 Paris Terrorist Attacks

**DOI:** 10.1101/2025.10.08.25337544

**Authors:** Alain Brunet, Isabelle Durand-Zaleski, Redwan Maatoug, Lina El-Houari, Mélanie Voyer, Nathalie Girault, Khalid Kalalou, Laurent Gugenheim, Nathalie Dzierzynski, Louis Jehel, Maud Rotharmel, Farah Hodeib, Alexis Bourla, Yohan Laverre, Isis Hanafy, Emmanuelle Castaigne, Anaël Ayrolles, Bernard Marc, Macarena Cuenca, Patrice Louville, Virginie Buisse, François Ducrocq, Marie-Odile Krebs, Dominique Januel, Stéphane Mouchabac, Olivier Guillin, Guillaume Vaiva, Ounsa Zia, Anne Bissery, Gaëlle Abgrall, Michel Benoit, Alicia Le Bras, Nematollah Jaafari, Ciara Treacy, Candice Estellat, Bruno Millet

## Abstract

**Background:** In response to the Bataclan terrorist attacks, the deadliest on French soil since World War II, the Paris Mémoire Vive (PARIS MEM) project was launched to expand the public hospital network’s capacity to treat trauma-related disorders. Reconsolidation Therapy (RT), a brief evidence-based intervention for post-traumatic stress disorder (PTSD), was selected for rapid implementation across hospital sites.

**Objective:** To evaluate the feasibility, effectiveness, and cost-utility of implementing RT to enhance public hospital treatment capacity after mass trauma exposure.

**Method:** A two-arm, multicentric, preference-based clinical trial and economic evaluation compared RT to treatment-as-usual (TAU) in 332 adults, mostly with PTSD. RT involves recalling trauma under propranolol during six weekly 25-minute sessions. Feasibility endpoints included the number of hospital staff trained and the proportion of participants choosing RT. The primary effectiveness endpoint was the delta scores in PTSD symptoms from baseline to Week 52. Cost-utility was evaluated using incremental cost-effectiveness ratios and quality-adjusted life years (QALYs).

**Results:** Across hospital sites, 160 therapists were trained in two days, and more participants opted for RT (n = 262) over TAU (n = 70), supporting feasibility endpoints. After one year, mean PTSD symptom scores decreased by 38.14 points (SD = 0.10) in RT and 35.02 (SD = 1.68) in TAU (p = .297), indexing significant improvement in both groups. At Week 7, RT showed faster initial recovery (difference = −5.11; p = .041). RT had a 55.4% probability of being cheaper and more effective than TAU (8.4%), with estimated savings of 27150€ per QALY. Annual sick-leave costs were lower for RT (4147€; 95% CI = 3 394–5 012) than TAU (7 386€; 95% CI = 5 416–10 340; p = .01).

**Conclusions:** Following massive trauma exposure, training mental health staff over two days in providing efficient, evidence-based, cost-effective PTSD treatment is achievable. Findings await replication.

**Trial Registration:** NCT02789982.

**Highlights:**

- It is feasible to train a large cohort of therapists in the aftermath of mass trauma and enhance the treatment capacities of institutions.
- Reconsolidation Therapy worked faster than treatment as usual in the alleviation of traumatic stress symptoms.
- Reconsolidation Therapy emerges as a cost-effective treatment, being both cheaper and more effective in 55% of cases.

---

The *Paris Mémoire Vive* (PARIS MEM) project was initiated in the wake of the Bataclan terrorist attacks, the deadliest event on French soil since WWII (Le Monde, 2015/11/13). Thousands of civilians were either victim or witness to one of five attacks that took place simultaneously in Paris on that day (Carli et al., 2017). In the subsequent days, public safety and hospital personnel were overwhelmed by a series of emergencies. To compound this state of affairs, it was anticipated that similar attacks might reoccur, potentially leading to a large, unmanageable influx of individuals seeking treatment for traumatic stress. Confronted with this scenario, the *Assistance Publique - Hôpitaux de Paris* (AP-HP) hospital network urgently sought ways to enhance the skills of its personnel and its treatment capacities.

Many evidence-based therapies for posttraumatic stress disorder (PTSD) are complex procedures that cannot be learned quickly (Cahill et al., 2009; Kirkpatrick and Heller, 2014). Although 2/3 of patients benefit from extinction-based therapies, 50% relapse within one year according to a well-known meta-analysis (e.g., Bradley et al., 2005). Similarly, while considered effective, psychotropic drugs have side effects that lead up to 32% of patients discontinuing their use, reducing overall efficiency (Bisson, 2007; Ipser & Stein, 2012; National Institute for Health and Care Excellence, 2018). Narrative exposure therapy (NET) is another trauma-focused intervention that involves constructing a coherent narrative to contextualize traumatic memories (Neuner et al., 2011). Whilst this modality offers a structured yet brief format adaptable to diverse and low-resource settings (Lely et al., 2019), NET nonetheless shares the same limitations as other extinction-based therapies and remains prone to relapse.

Reconsolidation Therapy (RT) is among the novel, evidence-based treatments for PTSD that can be taught quickly to clinicians (Brunet et al., 2008, 2018; Mallet et al., 2022). RT involves the delivery of six protocolized 25 min weekly session. During those sessions, patients briefly recall a traumatic event with a clinician 1 hour after ingesting a dose of oral propranolol, a ß-blocker. Propranolol decreases the aversiveness of the trauma recall and is a well-established memory reconsolidation blocker, subsequently decreasing the strength of the trauma memory (Brunet et al., 2008). Controlled trials support its efficacy (Brunet et al., 2018), especially for the more severe PTSD cases (Roullet et al., 2021). Three meta-analyses support its efficacy (Lonergan et al., 2013; Pigeon et al., 2022; Walsh et al., 2018), while two others are inconclusive (Astill Wright et al., 2021; Raut et al., 2022). Reconsolidation impairment, as a technique, is also considered less prone to relapse than extinction (Kindt et al., 2014).

We assessed the (i) feasibility of implementing an evidence-based PTSD treatment in a real-world setting in the aftermath of a series of traumatic event, (ii) its real-world clinical effectiveness, and (iii) its cost-utility as compared to the locally available treatments for PTSD: henceforth called treatment as usual (TAU). We hypothesized that clinicians working in real-life settings would learn and implement RT efficiently; at least 50% of the patient-participants would accept to receive time-limited RT vs. unlimited TAU; RT would be as effective as TAU but would confer a greater cost-benefit value because it presumably works faster.

## Methods and Materials

### Trial Design and Oversight

The complete protocol is published elsewhere (Brunet et al., 2019), but in brief, this is a 52 week non-randomized, preference-based, two-arm multicentric pragmatic clinical trial comparing the effectiveness and health economic impact of RT and TAU in a sample of trauma-exposed adults. Participants freely elected to receive either RT or TAU after discussing the matter with their doctor. For the RT group, weekly 25-minute treatment sessions were offered for six consecutive weeks vs. unlimited treatment sessions for TAU. Clinical measures were administered (single-blind) at baseline and at nine additional time points over the course of one year (weeks 1-7, 13, and 52). Cost-utility and feasibility data were gathered at the 13 and 52 Week time points. The project was approved by an ethics committee (CPP IdF6, n°14–16), the French data privacy commission (CNIL, n°916159), the EudraCT biomedical research agency (n°2016-000257-12) and conformed to the Helsinki guidelines. Clinicians received 16hr of manualized training developed at McGill’s Douglas Institute (Montréal, Canada) by the first author. A.B. trained small groups of clinicians and study personnel at sixteen French general psychiatry centers in Ile de France (12 centers), Lille, Fort-de-France (Martinique), Poitiers and Nice.

### Participants

The study criteria are fully described in Brunet et al. (2019). Briefly, treatment-seeking individuals were informed of the study upon their arrival for intake at the hospital. Written informed consent was obtained by the local study physician after the study personnel provided a detailed description of the trial. Included participants were required to have a current DSM-5 (American Psychiatric Association, 2013) diagnosis of a trauma and stressor-related disorder (mostly PTSD), a minimal Clinical Global Impression (CGI-S; Busner and Targum, 2007) severity rating of 3 (moderately ill), and a > 44 score on the PTSD Checklist (PCL-S;Ventureyra et al., 2002). The Mini International Neuropsychiatric Interview (M.I.N.I.; Sheehan et al., 1998) was used to ascertain diagnosis and comorbidity. Participants who opted for RT were seen by a physician and underwent a cardiogram. RT participants could not have a basal heart rate < 55 bpm, a systolic blood pressure < 100 mmHg, or a decrease of > 30% in vital signs 75 minutes after initial propranolol intake. Eligible participants were required to be on a stable medication regimen for > 2 months, if applicable. Drug-drug interactions were verified beforehand.

### Treatments

RT, as described in Brunet et al. (2018) consist in actively recalling one’s index event (i.e. worst/traumatic), using a specific memory reactivation protocol, under the influence of the ß-blocker propranolol, a well-established memory reconsolidation blocker. The protocol requires participants to attend a 25-min session with a therapist once a week, for 6 consecutive weeks. More specifically, 1 mg/kg of oral propranolol based on ideal body mass index is given on site 75 min before the session. Participants were asked to write (session 1) and read out loud (sessions 2–6) a 1–2 page narrative of their traumatic event. During subsequent sessions, participants were asked if they wish to update or modify their narrative to include new/more detailed information that bothered them during the previous week (Brunet et al., 2019). The RT workflow is presented in Appendix 1.

Participants enrolled in TAU received the PTSD treatment offered locally at the treatment center, whatever this treatment may be. In most centers, TAU took the form of an SSRI (most commonly, paroxetine 20–60 mg started at the first treatment visit), and/or psychotherapy such as cognitive-bahavior therapy (CBT), eye movement desensitization and reprocessing therapy (EMDR), supportive therapy or psychodynamic therapy.

### Endpoints

The PCL-5 was not validated in France at the time of study commencement, so the primary endpoint measure used the PCL-S (Blanchard et al., 1996). Secondary endpoint measures included the non-specific anxio-depressive symptom items of the Hopkins Symptom Checklist (HSCL-25; Ventevogel et al., 2007), the Clinical Global Impression Improvement scale (CGI-I), and the abridged World Health Organization Quality of Life (WHOQOL-BREF) scale (Whoqol Group, 1998). For the cost-utility feasibility endpoints, the French population’s utility-value set for each score of the EQ-5D-5L measure was used (Andrade et al., 2020; Luo et al., 2013). All measures were administered at baseline, Week 7 (i.e., one week after RT treatment completion), 13 and 52, except for the PCL-S, which was also administered weekly at Week 1-6.

### Statistical Analyses

Power was calculated *a priori* as a compromise between feasibility of recruitment and precision of the 95% confidence interval, based on a PCL-S within-group mean delta score of 25. Considering a standard deviation range of 15–25 (from Brunet et al., 2018) and a 1:1 group allocation ratio, 400 participants would lead to a 95%CI width of 4–7 points on the PCL-S. Analyses were conducted on the modified intention-to-treat (mITT) sample, involving all participants who had received at least one treatment session (Brody, 2016). Linear interpolation was used for the PCL-S missing data at Week 52 if the score was obtained beyond that time. Missing data was handled using multiple imputations via chained equation using 10 datasets. Analyses were weighted by propensity scores according to the inverse probability treatment weighting method with stabilized weights (Xu et al., 2010).

#### Clinical effectiveness

The primary clinical endpoint was the delta PTSD symptom scores from baseline to Week 52. Change from baseline PCL-S mean scores and 95%CIs for Weeks 7, 13 and 52 were estimated in both groups and compared using linear regression models adjusted according to PCL-S baseline scores. To validate the imputed data analyses, sensitivity analyses were performed on the per protocol sample, as well as using a mixed modelling approach that included repeated measures to impute missing data. Change from baseline PCL-S mean scores and 95%CIs for each of the first six treatment visits were estimated on an exploratory basis. A linear mixed-effects model with random intercept and slope was performed to describe the evolution of PCL-S change scores from baseline to Week 52 and their corresponding between-group differences. Those with a PCL-S score below the probable PTSD cut-off score of 44 points at week 52 were classified as treatment responders. Between-group comparisons of CGI-I involved linear mixed-effects modeling, adjusted according to baseline scores. Changes from baseline in anxio-depressive HSCL-25 scores at Weeks 7, 13 and 52 involved ordinal regressions. Analyses were performed using a two-sided alpha of 5%, with *R*-4.1.0 (R Foundation for Statistical Computing). No correction for multiple testing was used.

#### Feasibility

The number of clinicians trained in RT and the participants’ preference for RT or TAU were documented. Protocol adherence by healthcare professionals and treatment adherence were monitored through a list of indicators and was considered very good (Appendix 2). Serious adverse effects, adverse events, drop-outs, treatment responder rates and days of medical leave-of-absence were described and compared between groups.

#### Cost-utility

We estimated the cost-utility of RT vs. TAU from a societal viewpoint (i.e., healthcare and sick leave costs) with a 1-year time horizon (no discounting). Unit costs are presented in Appendix 3. In each group the median, mean and bootstrapped 95%CIs were computed for the cost estimates. Effectiveness was estimated using QALYs. The difference in QALYs was represented as the between-group difference in the plot area between the utility curves. The incremental cost-utility ratio was defined as the difference in mean total costs divided by the difference in mean total QALYs. The joint comparison of costs and effects was explored by non-parametric bootstrapping with 1 000 resamples represented on the cost-utility plane (Tehard et al., 2020).

## Results

### Feasibility

One hundred fifty clinicians and research staff were trained over two days in RT across 15 hospitals over a 10-week period (2016). Ten more clinicians were trained over two days in the Fort-de-France hospital at a later date (06/2017).

From 05/2016 to 01/2019, 401 participants were screened and 347 were included in the study: 78% of them selected RT (Figure 1) and 22% TAU. As shown in Figure 1, 15 participants were lost to follow-up prior to their first treatment visit, thus yielding a mITT sample of 332 participants (RT group = 262; TAU group = 70). The 1-year attrition rate in mITT was 26% (95%CI 21 - 32) for RT and 41% (95%CI 31 - 53) for TAU (*p* = .021).

**Figure 1.**
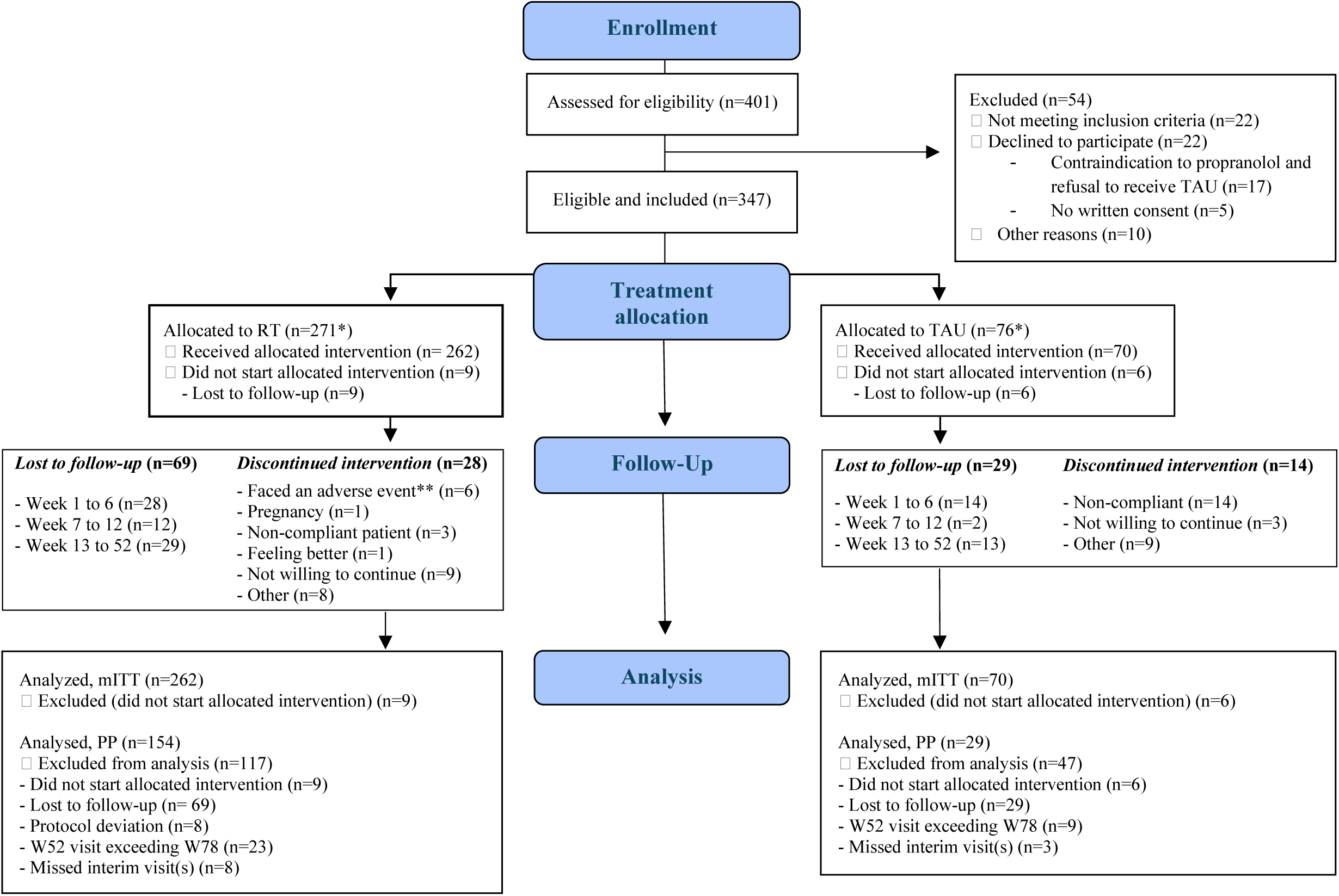
Study Flow Chart. W: week. RT: Reconsolidation Therapy. TAU: Treatment-as-usual. mITT: modified Intention to Treat population. PP: Per Protocol population. *Two patient-participants chose RT but were eventually allocated to TAU, while one patient-participant chose TAU but was eventually allocated to RT. These patient-participants were then lost to follow-up and therefore are not in the PP population. **Adverse events were anxiety, visual disturbances and feeling dizzy, peripheral vasoconstriction in the hands and feet, fever with myalgia and arthralgia, and major insomnia.

In terms of fidelity, 67% of the RT sessions were conducted one week apart, as planned. Vacations, schedule conflicts, or illness affected the other 33%. This statistic was not calculated for TAU since it has no predefined duration. For RT, 85% of the sessions were conducted 60-120 minutes post-propranolol, as recommended (Brunet et al., 2018). In the RT group, five adverse events led to treatment interruption for five individuals: anxiety, visual disturbances and feeling dizzy, peripheral vasoconstriction in the hands/feet, fever with myalgia and arthralgia (Figure 1). There were no adverse events in the TAU group.

Table 1 presents the socio-demographic variables. After imputation and weighting, the two groups did not differ, except for age, which could not be incorporated in the propensity score model (see Appendix 4) without unbalancing the CGI-I improvement variable. Anxious (66%) and depressive (78%) comorbidities were common across groups. There was no between-group difference in baseline illness severity (CGI-S): most participants were rated as *markedly* (63.2% vs. 65.7%) or *severely ill* (22.6% vs. 18.6%) in RT vs. TAU, respectively (*p* = .798).

**Table 1.** Socio-demographic Characteristics of the Samples. Multiple imputations generated 10 samples using the following variables: Symptom score (PCL-S), Clinical Global Impression (CGI) severity score, sex, trauma type, time of traumatic event, repeated trauma, study site, visit number, treatment group, concurrent treatment, non-psychiatric comorbidity, marital status, work status, educational level, mother tongue, depressive disorder, anxiety disorder, alcohol use disorder, obsessive-compulsive disorder. After imputation, propensity scores were computed, including for baseline PCL-S, baseline CGI (dichotomized in 4– 5 vs. 6–7), sex, trauma type, repeated trauma, concurrent treatment, non-psychiatric comorbidity, marital status, work status, educational level, mother tongue, depressive disorder, anxiety disorder, alcohol use disorder, obsessive-compulsive disorder. ASD (absolute standardized difference): An ASD < 0·1 means that the groups are balanced. TAU: Treatment-as-usual. RT: Reconsolidation Therapy. If patient-participants experienced multiple traumatic events, the most recent one was selected as the Index event. *p*-values are from Student’s *t*-test or Wilcoxon rank sum for continuous variables, and Chi-2 tests or Fisher exact tests for categorical variables, as appropriate.

| Characteristic | Raw Data |  |  |  |  | Imputed, Weighted Data |  |  |
| --- | --- | --- | --- | --- | --- | --- | --- | --- |
|  | n | TAU | n | RT | p-value | TAU (n=70) | RT (n=262) | ASD |
|  |  | *M (SD)* |  | *M (SD)* |  | *M (SD)* | *M (SD)* |  |
| Baseline PCL-S score | 69 | 64.24 (9.74) | 260 | 63.19 (11.22) | 0.722 | 64.91 (14.2) | 65.24 (3.62) | 0.02 |
| Age : M(SD) | 70 | 44.25(12.64) | 262 | 40.64 (12.89) | 0.034 | 44.66 (11.38) | 40.71(12.89) | 0.32 |
| Sex: |  | *n (%)* |  | *n (%)* |  | *%* | *%* |  |
| Female | 70 | 45 (64.29) | 262 | 190 (71.43) | 0.264 | 70.37 | 69.47 | -0.01 |
| Male |  | 25 (3.71) |  | 72 (28.57) |  | 29.63 | 30.53 | 0.01 |
| Work status: not working | 70 | 15 (21.43) | 262 | 66 (24.81) | 0.556 | 19.80 | 23.47 | 0.04 |
| Marital status: single | 70 | 36 (51.43) | 262 | 131 (49.25) | 0.745 | 47.73 | 49.30 | 0.02 |
| Mother tongue: French | 70 | 58 (82.86) | 262 | 239 (89.85) | 0.104 | 87.07 | 87.77 | 0.01 |
| Educational: junior high-school or + | 70 | 66 (94.29) | 262 | 260 (97.74) | 0.226 | 96.42 | 96.63 | 0.00 |
| Non-psychiatric comorbidity | 70 | 126 (47.37) | 262 | 42 (60.00) | 0.060 | 51.2 | 50.13 | -0.01 |
| Trauma type: terrorist attack | 70 | 75 (28.20) | 262 | 14 (20.00) | 0.167 | 28.42 | 27.03 | -0.01 |
| Repeated trauma | 69 | 13 (18.84) | 260 | 44 (16.67) | 0.669 | 14.15 | 16.78 | 0.03 |
| Concurrent treatment | 70 | 30 (42.86) | 260 | 68 (25.76) | 0.005 | 29.30 | 29.31 | 0.00 |
| Depressive disorder: |  |  |  |  |  |  |  |  |
| Current, isolated |  | 23 (32.86) |  | 81 (30.92) |  | 31.75 | 31.47 | 0.00 |
| Current, recurring |  | 23 (32.86) |  | 85 (32.82) | 0.902 | 34.26 | 32.88 | -0.01 |
| Past (not recurring) | 70 | 8 (11.43) | 261 | 38 (14.50) |  | 10.36 | 13.59 | 0.03 |
| None |  | 16 (22.86) |  | 57 (21.76) |  | 23.62 | 22.05 | -0.02 |
| Anxiety disorder: |  |  |  |  |  |  |  |  |
| Performance only |  | 31 (44.29) |  | 86 (33.21) |  | 32.54 | 35.45 | 0.03 |
| Full-fledged | 70 | 17 (24.29) | 261 | 85 (32.82) | 0.170 | 33.17 | 31.05 | -0.02 |
| None |  | 22 (31.43) |  | 90 (33.97) |  | 34.29 | 33.51 | -0.01 |
| Obsessive-compulsive dis. | 70 | 1 (1.43) | 261 | 22 (8.27) | 0.058 | 4.73 | 6.92 | 0.02 |
| Alcohol use disorder | 70 | 1 (1.43) | 261 | 17 (6.39) | 0.137 | 5.93 | 5.43 | 0.00 |
| Baseline CGI: severely ill or amongst most severely ill | 70 | 13 (18.57) | 261 | 62 (23.75) | 0.324 | 26.13 | 22.93 | -0.03 |

### Clinical Effectiveness

As shown in Figure 2, the PTSD symptom mean change scores did not significantly differ between RT (a 38.14-point improvement, 95%CI 36.50 – 39.77) and TAU (35.02, 95%CI 31.73 – 38.30) at Week 52 (*p* = .297), nor was there a significant between group-difference at Week 13 (*p* = .157). However, the Week 7 between-group difference was significant (*p* = .041), with a mean improvement of 33.40 points (95%CI 32.37 – 34.42) for the RT group versus 28.29 (95%CI 24.26 – 32.32) for the TAU group, a mean difference of 5.11 points on the PCL-S. A longitudinal analysis of the change in PCL-S scores indicated that there was a more rapid symptomatic improvement for the RT vs. TAU groups (*p < 0.001*). Data representing mean PTSD symptom improvement at each visit can be found in the supplemental material.

**Figure 2.**
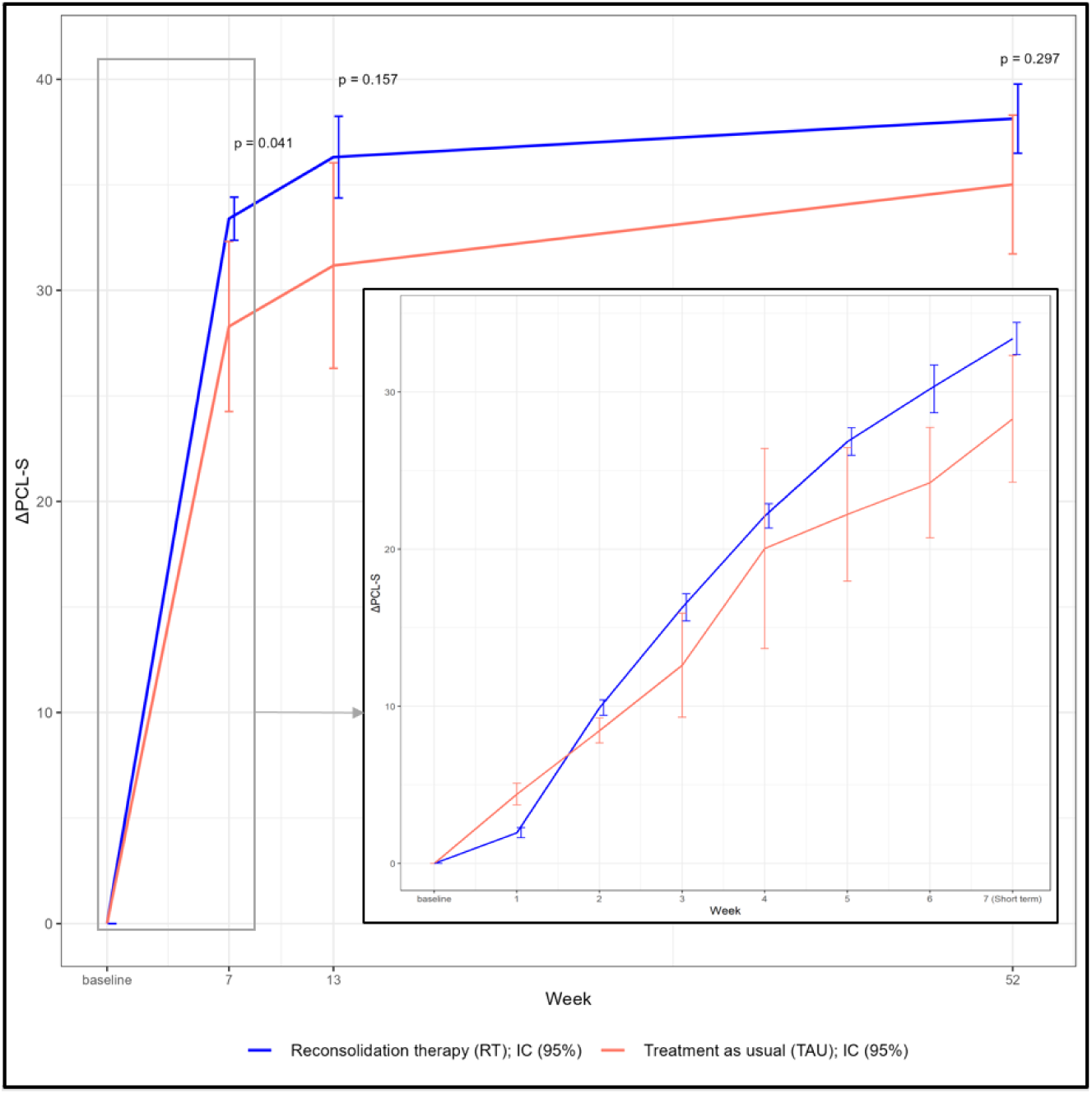
Self-rated PTSD Symptom Mean Improvement Score Over Time as a Function of Group. The *p*-values were obtained using regression models imputed and weighted on the propensity score and adjusted on group and the baseline value of the PTSD Symptom (PCL-S) scores. The confidence intervals (95%CI) do not overlap at Weeks 6 and 7, indicating a significant between-group difference, favoring RT vs. TAU. The *p*-values at Weeks 13 and 52, indicate that the between-group score differences are not statistically significant.

#### Sensitivity analyses

Analyses of clinical data using an imputed mixed modelling approach with repeated measures confirmed that RT was more effective than TAU at Week 7 (*p* = .048), and that there were no between-group differences at Week 13 (*p* = .266) and 52 (*p* = .123). In the per protocol analyses, these differences approached but did not reach statistical significance at each timepoint (data not shown).

#### Secondary clinical results

There were no between-group differences on the improvement scale of the CGI at Weeks 7 (*p* = .909), 13 (*p* = .362) or 52 (*p* = .877). The combined percentages of the CGI-I ratings of *Much improved* and *Very much improved* for the RT and TAU groups were: 55% vs. 52% for Week 7, 59% vs. 51% for Week 13, and 70% vs. 70% for Week 52 (Appendix 5), respectively.

We next examined treatment effects as per the non-specific anxio-depressive HSCL-25 total scores. Improvements from baseline to Week 52 were significant (*p* < 0.001) for both groups (RT: 0.69 ±0.04; TAU: 0.64 ±0.06). No between-group difference emerged for the HSCL-25 at Week 52 (*0.05*±0.*07, p=.615*) or at any other timepoint (Appendix 6).

#### Treatment Responders

The rate of treatment responders (i.e., PCL-S < 44) was higher in the RT group than the TAU group at all time points: Week 7 (RT: 58.1%, 95%CI 53.8 – 62.7; TAU: 46.8%, 95%CI 38.4 – 55.1, *p =* .*048*), Week 13 (RT: 66.1%, 95%CI 62.3 – 69.9; TAU: 52.8%, 95%CI 41.7 – 63.9, *p =* .*026*) and Week 52 (RT: 77.8%, 95%CI 74.0 – 81.6, TAU: 70.5%, 95%CI 63.4 – 77.6, *p =* .*030*).

### Cost-utility Analyses

The difference in mean direct annual costs incurred with RT (2 746€, 95%CI 2 272 - 3 276) versus TAU (3 289€, 95%CI 2 191 - 5 598) were non-significant (*p* = .47, see Table 2). Mean QALYs were 0.809 (95%CI: 0.791 – 0.831) and 0.807 (95%CI: 0.748 – 0.839) for the RT and TAU groups, respectively (*p* = .94). RT yielded a significantly greater cost effectiveness compared to TAU, with a point estimate of 271 500€ saved per additional QALY. The annual sick leave costs for the RT group were 4 147€ (95%CI: 3 394 – 5 012) whilst TAU was 7 386€ (95%CI: 5 416 – 10 340), a significant difference (*p* = .01). RT had a 55.4% probability of being both more effective and less costly than TAU (Figure 3).

**Figure 3.**
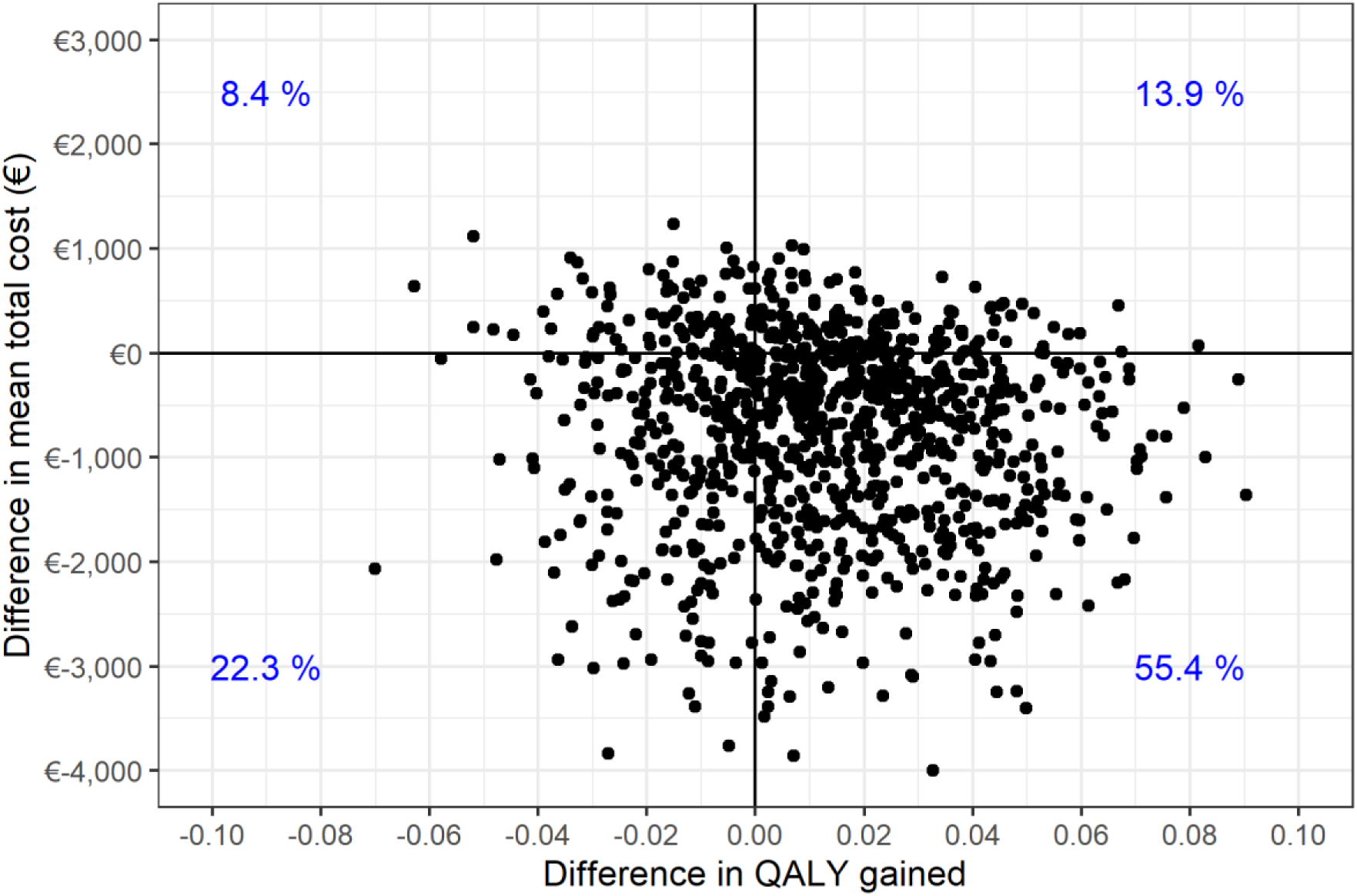
Difference in QALY for Reconsolidation Therapy versus Treatment As Usual. Bootstrap of the Distribution of the 1000 ICER (in €/QALY) at One Year – Societal Perspective, Imputed and Weighted by Propensity Scores

**Table 2.** Cost of Treatment Over a Year. Average Cost Per Patient-participant at One-Year according to treatment modality

|  | Reconsolidation<br>Therapy<br>(N = 262) | Treatment<br>as Usual<br>(N = 70) | P-value |
| --- | --- | --- | --- |
| Reconsolidation/standard therapy | 349 [339 ; 358] | 326 [297 ; 345] | 0.09 |
| Medical consultations | 691 [616 ; 775] | 838 [681 ; 1,116] | 0.21 |
| • *Psychiatrist* | 328 [270 ; 397] | 335 [220 ; 502] | 0.93 |
| • *Others* | 363 [323 ; 402] | 503 [396 ; 667] | 0.03 |
| Paramedical consultations | 199 [157 ; 246] | 256 [146 ; 382] | 0.39 |
| Psychologists | 123 [ 80 ; 176] | 223 [105 ; 397] | 0.14 |
| Alternative and complementary medicine | 78 [ 59 ; 99] | 159 [ 67 ; 278] | 0.08 |
| Drugs | 187 [140 ; 244] | 251 [194 ; 336] | 0.16 |
| Hospitalisations | 1 067 [616 ; 1 550] | 1 165 [312 ; 41 932] | 0.88 |
| • *In medicine, surgery or obstetrics* | 354 [240 ; 483] | 214 [100 ; 613] | 0.18 |
| • *In psychiatry* | 713 [315 ; 3 646] | 951 [124 ; 45 402] | 0.68 |
| Call to emergency services | 2 [ 1 ; 3] | 2 [ 1 ; 5] | 0.58 |
| Emergency department visits | 4 [ 2 ; 6] | 6 [ 2 ; 16] | 0.38 |
| Ambulance transport | 12 [ 6 ; 18] | 14 [ 5 ; 29] | 0.68 |
| Social worker | 34 [ 23 ; 47] | 49 [ 23 ; 69] | 0.38 |
| TOTAL | 2 746 [2 272 ; 3 276] | 3 289 [2 191 ; 5 598] | 0.47 |

## Discussion

### Clinical Effectiveness

In this trial, as planned, both RT and TAU showed similar reductions in symptoms at 52 weeks. However, given that manualized RT is delivered over 6 weeks and many treatments subsumed under TAU (e.g., SSRIs, EMDR, and CBT) typically require 6 to 12 weeks to show a response (Roberts et al., 2016), it was considered reasonable to analyze treatment response across groups at Week 7 and 13 to ensure a fair comparison. RT produced greater treatment responses than TAU at Week 7, with a between-group difference slightly over 5 points on the study’s PTSD symptom measure. This score magnitude is considered both clinically meaningful and reliable (Marx et al., 2022).

### Feasibility

This trial aimed to provide real-world evidence on the feasibility of quickly teaching and implementing in an emergency situation an evidence-based treatment into routine practice following large-scale terrorist attacks, to enhance preparedness in a public hospital network. This pragmatic approach also offered a practical extension of a double-blind RCT which evaluated RT vs. placebo in Canada just prior to current study commencement (Brunet et al., 2018). Indeed, the present study design captures the complex, time-sensitive, and heterogenous nature of PTSD treatment in the immediate aftermath of such a chaotic event, where other methodologies may fall short. Nonetheless, opting to teach RT seemed like a bold idea: Could the treatment really be taught in 2 days? Would patient-participants feel short-changed by receiving only six, 25-minute treatment sessions instead of unlimited TAU? Would clinicians who are used to providing their preferred treatment agree to give it a try? In this trial, both participants and clinicians considered RT an attractive treatment option, as evidenced by the training of 160 clinicians, and by the 3:1 uptake ratio in favor of RT by patients, a ratio much higher than the 1:1 ratio we had hypothesized. This data suggests that, in the aftermath of a large-scale traumatic event, most clinicians and treatment-seeking participants are supportive of trying novel and rapid evidence-based interventions, indicating a strong interest for RT.

### Cost-utility

Although RT equaled or outperformed TAU on the clinical endpoints over the 1-year study period, it achieved these outcomes with fewer resources, requiring only six 25-minute sessions delivered by clinicians who received 16-hours of brief specialized training. Importantly, the economic analysis revealed that RT had a 55% probability of dominance compared to 8% for TAU, underscoring RT’s potential as a scalable, cost-efficient intervention. When factoring in the cost of sick leave compensation, RT fared more favorably than TAU over the one-year follow-up period.

### Study limitations

The main limitations of non-randomized pragmatic trials pertain to their internal validity. Although propensity score matching was used to reduce confounding by equating treatment groups based on dozens of covariates, this equating is dependent on the covariates that are included in the matching. As such, other unmeasured variables may have influenced our results. The lack of double-blind assessments is another source of potential bias in the current study. However, given nearly all psychotherapy trials are conducted in a single blind manner, for obvious reasons: the patients inevitably know what treatment they are receiving.

Although a study spanning one year is longer than what is commonly found in many trials (Ward et al., 2025), a more complete representation of the therapeutic effects examined with regards to mental health and resource utilization could be obtained from a longer study. Moreover, the administration of service use questionnaires, rather than actual service utilization, can introduce memory bias and lead to an underestimation of costs. Whilst a difference of one QALY may appear modest, it becomes more meaningful when considered alongside the finding that RT was likely to be both more effective and less costly than other treatment methods, even when evaluated using the EQ-5D-5L, a very conservative measure for economic evaluations. Finally, RT was compared to a heterogeneous set of therapeutic practices (TAU) for which expertise and treatment adherence was not monitored. It remains unknown how RT would perform in a direct comparison with any of these alternate therapeutic methods in a randomized controlled trial. This paper should not be taken as evidence that RT is superior in effectiveness to other PTSD treatment methods. It was however found to be very efficient, obtaining good outcomes while using few clinical resources.

## Concluding Remarks

In this carefully crafted study, RT emerged as the preferred and generally more cost-effective treatment option compared to TAU following the Bataclan terrorist attacks, showing greater PTSD symptom reduction at 7 weeks and comparable improvements at the 1 year mark. RT is appealing because it is easily taught and integrated into routine practice, offering a treatment option that may enable a greater number of PTSD patients to be successfully treated in less time compared to other therapies. This easily implemented treatment may prove particularly valuable when managing trauma victims in the aftermath of large-scale traumatic events, such as wars or the growing frequency of natural disasters. These promising findings add to the mounting evidence in favor of RT and call for replication in larger randomized controlled trials across various traumatic event settings.

## Data Availability

The datasets used and/or analyzed during the current study belong to the AP-HP (Assistance Publique-Hopitaux de Paris) research directorate and will be available by reasonable request to the corresponding author.

## Authors’ contributions

Author names and abbreviations: Alain Brunet, (A.B.), Isabelle Durand-Zalesky, (I.D.Z.), Redwan Maatoug, (R.M.), Lina El-Houari, (L.E.H.), Mélanie Voyer, (M.V.), Nathalie Girault, (N.G.), Khalil Kalalou (K.K.), Laurent Gugenheim, (L.G.), Nathalie Dzierzynski, (N.D.), Louis Jehel, (L.J.), Maud Rotharmel, (M.R.), Farah Hodeib, (F.H.), Alexis Bourla, (A.B. Bourla), Yohan Laverre, (Y.L.), Isis Hanafy, (I.H.), Emmanuelle Castaigne, (E.C.), Anaël Ayrolles, (A.A.), Bernard Marc (B.M.), Macarena Cuenca (M.C.), Patrice Louville (P.L.), Virginie Buisse (V.B.), François Ducrocq, (F.D.), Marie-Odile Krebs, (M.K.), Dominique Januel, (D.J.), Stéphane Mouchabac, (S.M.), Olivier Guillin, (O.G.), Guillaume Vaiva, (G.V.), Ounsa Zia (O.Z.), Anne Bissery, (A.B. Bissery), Gaëlle Abgrall, (G.A.), Michel Benoit, (M.B.), Nematollah Jaafari, (N.J.), Alicia Le Bras (A.L.), Ciara Treacy (C.T.), Candice Estellat, (C.E.), & Bruno Millet, (B.M.).

*Conceptualization:* A.B., I.D.Z., & B.M.

*Data curation:* A.B, I.D.Z., R.M., L.E.H., A.L., C.E., & B.M.

*Formal analysis:* A.B, I.D.Z., R.M., L.E.H., A.L., C.E., & B.M.

*Investigation:* A.B., R.M., M.V., N.G., K.K., L.G., N.D., L.J., M.R., F.H., A.B. (Bourla), Y.L., I.H., E.C., A.A., B.M. (Marc), M.C., P.L., V.B., F.D., M.K., D.J., S.M., O.G., G.V., O.Z., A.B. (Bissery), G.A., M.B., N.J., & B.M.

*Methodology:* A.B., I.D.Z., & B.M.

*Project administration:* A.B., I.D.Z., & B.M.

*Supervision:* A.B., I.D.Z., & B.M.

*Validation:* A.B., I.D.Z., R.M., L.E.H., M.V., N.G., K.K., L.G., N.D., L.J., M.R., F.H., A.B. (Bourla), Y.L., I.H., E.C., A.A., B.M. (Marc), M.C., P.L., V.B., F.D., M.K., D.J., S.M., O.G., G.V., O.Z., A.B. (Bissery), G.A., M.B., A.L., N.J., C.E. & B.M.

*Visualization:* A.B., I.D.Z., R.M., L.E.H., A.L., C.E. & B.M.

*Writing – original draft:* A.B., I.D.Z., R.M., L.E.H., A.L., C.E. & B.M.

*Writing – review & editing:* A.B., I.D.Z., R.M., L.E.H., M.V., N.G., K.K., L.G., N.D., L.J., M.R., F.H., A.B.

(Bourla), Y.L., I.H., E.C., A.A., B.M. (Marc), M.C., P.L., V.B., F.D., M.K., D.J., S.M., O.G., G.V., O.Z., A.B.

(Bissery), G.A., M.B., A.L., N.J., C.T., C.E. & B.M.

## Acknowledgements

We thank the study sites and their principal investigators (in parentheses) : Hôpital de la Pitié-Salpêtrière de Paris (Girault); EPS de Ville Evrard (Januel), Hôpital Tenon de Paris (Abgrall), Hôpital Corentin-Celton (Louville, Devulder), Centre hospitalier régional universitaire de Lille (Ducrocq, Vaiva), Hôpital Pasteur de Nice (Benoit), Centre Hospitalier de Marne-la-Vallée (Marc), Hôpital Sainte-Anne de Paris (Krebs), Hôpital Le Kremlin-Bicêtre de Paris (Castaigne), Hôpital Saint-Antoine de Paris (Mouchabac), Centre hospitalier Henri-Laborit de Poitiers (Jaafari), Hôpital Esquirol de St-Maurice (Cabié), Centre hospitalier universitaire du Rouvray (Guillin), Hôpital Henri-Mondor de Créteil (Botero), and Centre Hospitalier des Antilles Françaises, La Martinique (Jehel).

We thank the Deniker Foundation for its support. We thank Martin Hirsh, former AP-HP Director, who facilitated the launch of the project. We thank the personnel of the Pitié-Salpêtrière Clinical Research Unit, especially A. Mallet, L. Gambotti, and M. Prevost for conceptual input, coordination, study monitoring and data management, and the personnel of the AP-HP (Y. Vacher) for fulfilling regulatory tasks. We thank the anonymous panel who independently reviewed the study proposal for the AP-HP. We thank the *Fondation de l’AP-HP* and *MSD Avenir* for funding the project; neither were involved in the analysis of the study results or in their interpretation.

## Data Availability Statement

Data is available upon reasonable request from Candice Estellat.

## References

American Psychiatric Association, 2013. Diagnostic and statistical manual of mental disorders: DSM-5. American psychiatric association Washington, DC.

Andrade, L.F., Ludwig, K., Goni, J.M.R., Oppe, M., de Pouvourville, G., 2020. A French Value Set for the EQ-5D-5L. Pharmacoeconomics. 38, 413–425. 10.1007/s40273-019-00876-4.

Astill Wright, L., Horstmann, L., Holmes, E.A., Bisson, J.I., 2021. Consolidation/reconsolidation therapies for the prevention and treatment of PTSD and re-experiencing: a systematic review and meta-analysis. Translational Psychiatry. 11, 453. 10.1038/s41398-021-01570-w.

Bisson, J.I., 2007. Post-traumatic stress disorder. BMJ. 334, 789–793. 10.1136/bmj.39162.538553.80.

Blanchard, E.B., Jones-Alexander, J., Buckley, T.C., Forneris, C.A., 1996. Psychometric properties of the PTSD Checklist (PCL). Behaviour Research and Therapy. 34, 669–673. 10.1016/0005-7967(96)00033-2.

Bradley, R., Greene, J., Russ, E., Dutra, L., Westen, D., 2005. A Multidimensional Meta-Analysis of Psychotherapy for PTSD. American Journal of Psychiatry. 162, 214–227. 10.1176/appi.ajp.162.2.214.

Brody, T. (2016). Chapter 8 – Intent-to-Treat Analysis Versus Per Protocol Analysis. Clinical Trials (2^nd^ Edition). 10.1016/B978-0-12-804217-5.00008-4

Brunet, A., Ayrolles, A., Gambotti, L., Maatoug, R., Estellat, C., Descamps, M., Girault, N., Kalalou, K., Abgrall, G., Ducrocq, F., Vaiva, G., Jaafari, N., Krebs, M.O., Castaigne, E., Hanafy, I., Benoit, M., Mouchabac, S., Cabie, M.C., Guillin, O., Hodeib, F., Durand-Zaleski, I., Millet, B., 2019. Paris MEM: a study protocol for an effectiveness and efficiency trial on the treatment of traumatic stress in France after the 2015-16 terrorist attacks. BMC Psychiatry. 19, 351. 10.1186/s12888-019-2283-4.

Brunet, A., Orr, S.P., Tremblay, J., Robertson, K., Nader, K., Pitman, R.K., 2008. Effect of post-retrieval propranolol on psychophysiologic responding during subsequent script-driven traumatic imagery in post-traumatic stress disorder. Journal of Psychiatric Research. 42, 503–506. 10.1016/j.jpsychires.2007.05.006.

Brunet, A., Saumier, D., Liu, A., Streiner, D.L., Tremblay, J., Pitman, R.K., 2018. Reduction of PTSD Symptoms With Pre-Reactivation Propranolol Therapy: A Randomized Controlled Trial. American Journal of Psychiatry. 175, 427–433. 10.1176/appi.ajp.2017.17050481.

Busner, J., Targum, S.D., 2007. The clinical global impressions scale: applying a research tool in clinical practice. Psychiatry (Edgmont). 4, 28–37.

Cahill, S.P., Rothbaum, B.O., Resick, P.A., Follette, V.M., 2009. Cognitive-behavioral therapy for adults, In: Effective treatments for PTSD: Practice guidelines from the International Society for Traumatic Stress Studies, 2nd ed. The Guilford Press, New York, NY, US, pp. 139–222.

Carli, P., Pons, F., Levraut, J., Millet, B., Tourtier, J.P., Ludes, B., Lafont, A., Riou, B., 2017. The French emergency medical services after the Paris Nice terrorist attacks: what have we learnt? Lancet. 390, 2735–2738. 10.1016/s0140-6736(17)31590-8.

Ipser, J.C., Stein, D.J., 2012. Evidence-based pharmacotherapy of post-traumatic stress disorder (PTSD). International Journal of Neuropsychopharmacology. 15, 825–840. 10.1017/S1461145711001209.

Kindt, M., Soeter, M., & Sevenster, D. (2014). Disrupting reconsolidation of fear memory in humans by a noradrenergic β-blocker. Journal of Visualized Experiments. 94, 52151. 10.3791/52151

Kirkpatrick, H.A., Heller, G.M., 2014. Post-traumatic stress disorder: theory and treatment update. The International Journal of Psychiatry in Medicine. 47, 337–346. 10.2190/PM.47.4.h.

Lely, J. C. G., Smid, G. E., Jongedijk, R. A., W Knipscheer, J., & Kleber, R. J. (2019). The effectiveness of narrative exposure therapy: a review, meta-analysis and meta-regression analysis. European Journal of Psychotraumatology. 10, 1550344. 10.1080/20008198.2018.1550344

Lonergan, M.H., Olivera-Figueroa, L.A., Pitman, R.K., Brunet, A., 2013. Propranolol’s effects on the consolidation and reconsolidation of long-term emotional memory in healthy participants: a meta-analysis. Journal of Psychiatry and Neuroscience. 38, 222. 10.1503/jpn.120111.

Luo, N., Li, M., Chevalier, J., Lloyd, A., Herdman, M., 2013. A comparison of the scaling properties of the English, Spanish, French, and Chinese EQ-5D descriptive systems. Quality of Life Research. 22, 2237–2243. 10.1007/s11136-012-0342-0.

Mallet, C., Chick, C.F., Maatoug, R., Fossati, P., Brunet, A., Millet, B., 2022. Memory reconsolidation impairment using the beta-adrenergic receptor blocker propranolol reduces nightmare severity in patients with posttraumatic stress disorder: a preliminary study. Journal of Clinical Sleep Medicine. 18, 1847–1855. 10.5664/jcsm.10010.

Marx, B.P., Lee, D.J., Norman, S.B., Bovin, M.J., Sloan, D.M., Weathers, F.W., Keane, T.M., Schnurr, P.P., 2022. Reliable and clinically significant change in the clinician-administered PTSD Scale for DSM-5 and PTSD Checklist for DSM-5 among male veterans. Psychological Assessment. 34, 197–203. 10.1037/pas0001098.

National Institute for Health and Care Excellence, 2018. Post-traumatic stress disorder NG116. https://www.nice.org.uk/guidance/ng116.

Neuner, F., Elbert, T., & Schauer, M. (2011). *Narrative Exposure Therapy: A short-term intervention for traumatic stress disorders*. Hogrefe.

Pigeon, S., Lonergan, M., Rotondo, O., Pitman, R.K., Brunet, A., 2022. Impairing memory reconsolidation with propranolol in healthy and clinical samples: a meta-analysis. Journal of Psychiatry and Neuroscience. 47, E109–e122. 10.1503/jpn.210057.

Raut, S.B., Canales, J.J., Ravindran, M., Eri, R., Benedek, D.M., Ursano, R.J., Johnson, L.R., 2022. Effects of propranolol on the modification of trauma memory reconsolidation in PTSD patients: A systematic review and meta-analysis. Journal of Psychiatric Research. 150, 246–256. 10.1016/j.jpsychires.2022.03.045.

Roberts, N.P., Roberts, P.A., Jones, N., Bisson, J.I., 2016. Psychological therapies for post-traumatic stress disorder and comorbid substance use disorder. Cochrane Database of Systematic Reviews. 4, CD010204. 10.1002/14651858.CD010204.pub2.

Roullet, P., Vaiva, G., Very, E., Bourcier, A., Yrondi, A., Dupuch, L., Lamy, P., Thalamas, C., Jasse, L., El Hage, W., Birmes, P., 2021. Traumatic memory reactivation with or without propranolol for PTSD and comorbid depressive symptoms: a randomised clinical trial. Neuropsychopharmacology. 46, 1643–1649. 10.1038/s41386-021-00984-w

Sheehan, D.V., Lecrubier, Y., Sheehan, K.H., Amorim, P., Janavs, J., Weiller, E., Hergueta, T., Baker, R., Dunbar, G.C., 1998. The Mini-International Neuropsychiatric Interview (M.I.N.I.): the development and validation of a structured diagnostic psychiatric interview for DSM-IV and ICD-10. The Journal of Clinical Psychiatry. 59 Suppl 20, 22–57.

Tehard, B., Detournay, B., Borget, I., Roze, S., De Pouvourville, G., 2020. Value of a QALY for France: A New Approach to Propose Acceptable Reference Values. Value Health. 23, 985–993. 10.1016/j.jval.2020.04.001.

Ventevogel, P., De Vries, G., Scholte, W.F., Shinwari, N.R., Faiz, H., Nassery, R., van den Brink, W., Olff, M., 2007. Properties of the Hopkins Symptom Checklist-25 (HSCL-25) and the Self-Reporting Questionnaire (SRQ-20) as screening instruments used in primary care in Afghanistan. Social Psychiatry and Psychiatric Epidemiology. 42, 328–335. 10.1007/s00127-007-0161-8.

Ventureyra, V. A., Yao, S. N., Cottraux, J., Note, I., & De Mey-Guillard, C. (2002). The validation of the Posttraumatic Stress Disorder Checklist Scale in posttraumatic stress disorder and nonclinical subjects. Psychotherapy and Psychosomatics, 71, 47–53. 10.1159/000049343

Walsh, K.H., Das, R.K., Saladin, M.E., Kamboj, S.K., 2018. Modulation of naturalistic maladaptive memories using behavioural and pharmacological reconsolidation-interfering strategies: a systematic review and meta-analysis of clinical and ‘sub-clinical’ studies. Psychopharmacology. 235, 2507–2527. 10.1007/s00213-018-4983-8.

Ward, W., Haslam, A., & Prasad, V. (2025). Antidepressant Trial Duration Versus Duration of Real-World Use: A Systematic Analysis. The American Journal of Medicine, 138,, 1400–1407.10.1016/j.amjmed.2025.04.037

Whoqol Group, 1998. The World Health Organization Quality of Life Assessment (WHOQOL): development and general psychometric properties. Social Science & Medicine. 46, 1569–1585. 10.1016/S0277-9536(98)00009-4.

Xu, S., Ross, C., Raebel, M.A., Shetterly, S., Blanchette, C., Smith, D., 2010. Use of stabilized inverse propensity scores as weights to directly estimate relative risk and its confidence intervals. Value Health. 13, 273–277. 10.1111/j.1524-4733.2009.00671.x.

