## Appendix 1 for "Feasibility, Effectiveness, and Cost-utility of Implementing a Reconsolidation-Based Trauma Treatment in the Aftermath of the November 13 Paris Terrorist Attacks"

Workflow of Session 1-6 of Reconsolidation Therapy.

| Session 1 | | | | |
| --- | --- | --- | --- | --- |
| Step | Duration (min) | | | Professional involved |
| Greet participant | | 1 | Secretary | |
| Explain treatment, give snack | | 7 | Doctor, psychologist, or nurse | |
| Take vital signs | | 1 | Nurse | |
| Give propranolol | | 2 | Doctor, or nurse | |
| Write trauma narrative | | n/a | None | |
| Take vital signs | | 1 | Nurse | |
| Consultation | | 50 | Doctor, or psychologist | |
| Departure of participant | | 2 | Secretary | |
| Sessions 2-6 | | | | |
| Step | | Duration (min) | | Professional involved |
| Greet participant | | 1 | | Secretary |
| Give snack | | 5 | | Doctor, psychologist, or nurse |
| Give propranolol | | 2 | | Doctor, or nurse |
| Edit trauma narrative  (if needed) | | n/a | | None |
| Consultation | | 25 | | Doctor, or psychologist |
| Departure of participant | | 2 | | Secretary |
