## Appendix 2 for "Feasibility, Effectiveness, and Cost-utility of Implementing a Reconsolidation-Based Trauma Treatment in the Aftermath of the November 13 Paris Terrorist Attacks"

Treatment Adherence Monitoring Methodology for Reconsolidation Therapy.

Unless stated otherwise, RT treatment adherence was systematically monitored (100%) by the staff for all participants and clinicians using the following reliability checks: provision of a treatment rationale, getting a medical exam, determination of propranolol dosage, prescription of oral propranolol, assessment of basal vital signs (HR x2 and BP x2), evaluation of side-effect by the nurse (89%), monitoring time elapsed between propranolol ingestion (x6) and the beginning of the session (between 60-120 min.: 85%), verification of the trauma history and case report form by the treating clinician, undergoing a diagnostic interview, selection and confirmation of the *index* stressor to work on, meeting with a clinician (x2) conducting the clinical pre- and post-treatment assessments (CGI), completion of the weekly PTSD (x6) self-reported PTSD symptom measure (PCL-S), writing a 1-2 page trauma narrative (98%), use of 1^st^ person singular (99%), present tense for the trauma narrative (95%), inclusion in the narrative of physical sensations drawn from a list, confirmation that the participants read their trauma narrative (x6), identification by the participant to the therapist of the narrative’s *hot spot* (x6; 96%), updating (if needed) the trauma narrative at each subsequent visit (x5), prescribed number of treatment sessions (97%), number of days (7) between treatment each visit (66%), as well as failing to make use of any other empirically-validated treatment method (x6) for treating PTSD by the clinician during RT (e.g., cognitive restructuring, bilateral stimulations, interpretation, etc.). Treatment adherence was deemed excellent, expect for time elapsed between each visit: for the most part, very minor departures were noted which were immediately corrected via feedback to the clinicians.
