## Appendix 3 for "Feasibility, Effectiveness, and Cost-utility of Implementing a Reconsolidation-Based Trauma Treatment in the Aftermath of the November 13 Paris Terrorist Attacks"

Unit Costs (€).

|  | Healthcare system (producer’s) perspective | Health insurer’s perspective |
| --- | --- | --- |
| Medical professionnals |  |  |
| Anesthesist; reanimator | 45 | 26 |
| Surgeon | 51 | 26 |
| Dermatologist; infectiologist | 41 | 25 |
| Endocrinologist | 48 | 30 |
| Gastro-enterologist; hepatologist | 46 | 30 |
| General practitioner | 26 | 19 |
| Geriatrist | 38 | 26 |
| Gynecologist; obstetrician | 44 | 20 |
| Internist | 32 | 26 |
| Nephrologist | 43 | 36 |
| Neurologist | 64 | 47 |
| Ophtalmologist | 45 | 20 |
| Otorhinolaryngologist | 49 | 27 |
| Cardiologist | 53 | 42 |
| Pediatrist | 34 | 17 |
| Pneumologist | 38 | 28 |
| Psychiatrist | 60 | 35 |
| Rhumatologist | 45 | 28 |
| Stomatologist | 46 | 26 |
| Other professionnals |  |  |
| Dentist | 23 | 16 |
| Dietetician | 40 | 0 |
| Nurse | 9 | 8 |
| Kinesiotherapist; massage therapist | 18 | 13 |
| Orthophonist | 32 | 24 |
| Orthopedist | 24 | 15 |
| Podologist | 27 | 15 |
| Mid-wife | 30 | 29 |
| Psychologist | 60 | 0 |
| Complementary medicine |  |  |
| Bonesetters | 80 | 0 |
| Sophrologist | 50 | 0 |
| Acupuncturist | 52 | 0 |
| Naturopath; phytotherapist | 54 | 0 |
| Osteopath | 56 | 0 |
| Ambulance call | 6 | 0 |
| Emergency room visit | 78 | 55 |
| Transport ambulance (2-way) | 189 | 174 |
| Social worker | 50 | 0 |
