## Supplementary figures and images for "Feasibility, Effectiveness, and Cost-utility of Implementing a Reconsolidation-Based Trauma Treatment in the Aftermath of the November 13 Paris Terrorist Attacks"

### Appendix 4

Appendix 4

Balancing Covariates in the Study Groups Using Propensity Scores.

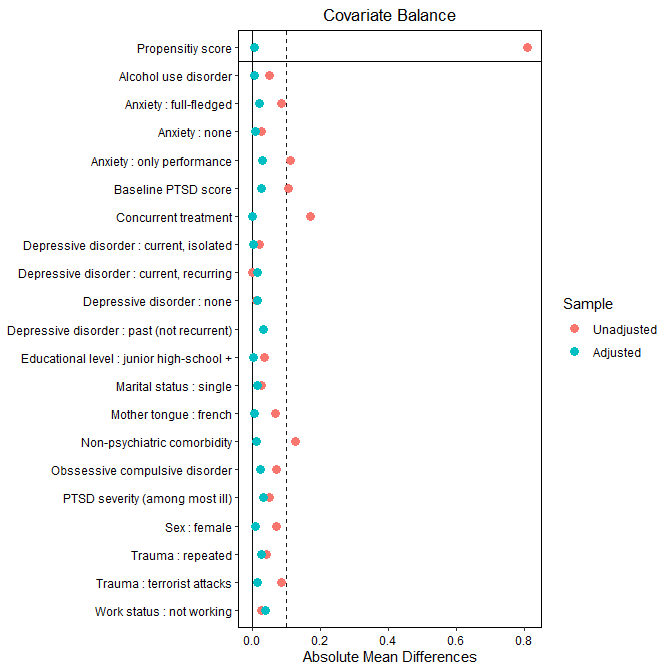
