## Appendix 5 for "Feasibility, Effectiveness, and Cost-utility of Implementing a Reconsolidation-Based Trauma Treatment in the Aftermath of the November 13 Paris Terrorist Attacks"

Clinical Global Improvement Ratings as a Function of Treatment Group: Raw vs. Imputed Weighted Data.

|  |  | Week 7 | | | Week 13 | | | Week 52 | | |
| --- | --- | --- | --- | --- | --- | --- | --- | --- | --- | --- |
| Raw  data  n (%) |  | TAU | RT | *p*-value* | TAU | RT | *p-*value* | TAU | RT | *p-*value* |
|  | Rating | *n*=48 | *n*=221 |  | *n*=46 | *n*=209 |  | *n*=39 | *n*=185 |  |
|  | Very much improved | 11 (23) | 40 (18) | 0.655 | 12 (26) | 61 (29%) | 0.582 | 9 (23) | 59 (32) | 0.274 |
|  | Much improved | 16 (33) | 86 (39) |  | 13 (28) | 67 (32%) |  | 16 (41) | 72 (39) |  |
|  | Slightly improved | 17 (36) | 71 (32) |  | 17 (37) | 63 (30%) |  | 11 (28) | 41 (22) |  |
|  | No  change | 4 (8) | 15 (7) |  | 4 (9) | 14 (7%) |  | 2 (5) | 9 (5) |  |
|  | Slightly worse | - | 9 (4) |  | - | 4 (2%) |  | - | 4 (2) |  |
|  | Much worse | - | - |  | - | - |  | 1 (3) | - |  |
|  | Very much  Worse | - | - |  | - | - |  | - | - |  |
|  |  | *n*=70 | *n*=262 |  | *n*=70 | *n*=262 |  | *n*=70 | *n*=262 |  |
| Imputed Weighted data  n (%) | Very much improved | 17 (24) | 45 (17) | 0.909 | 19 (27) | 73 (28) | 0.362 | 21 (30) | 81 (31) | 0.877 |
|  | Much improved | 20 (28) | 100 (38) |  | 17 (24) | 81 (31) |  | 27 (39) | 102 (39) |  |
|  | Slightly improved | 24 (35) | 86 (33) |  | 27 (39) | 79 (30) |  | 16 (24) | 55 (21) |  |
|  | No  change | 8 (11) | 21 (8) |  | 5 (7) | 18 (7) |  | 3 (4) | 16 (6) |  |
|  | Slightly worse | 1 (2) | 10 (4) |  | 2 (2) | 5 (2) |  | 1 (1) | 5 (2) |  |
|  | Much worse | - | - |  | - | 3 (1) |  | 2 (2) | 3 (1) |  |
|  | Very much  worse | - | - |  | - | - |  | - | - |  |

**p*-values were obtained using ordinal regression models using raw data and imputed data weighted on propensity score after multiple imputations. Percentages presented were rounded to the next unit.
