## Appendix 6 for "Feasibility, Effectiveness, and Cost-utility of Implementing a Reconsolidation-Based Trauma Treatment in the Aftermath of the November 13 Paris Terrorist Attacks"

Mean PCL-S and HSCL-25 Scores from Baseline to Week 7, 13 and 52 in Reconsolidation Therapy (RT) and Treatment-as-usual (TAU) Groups: Raw vs. Imputed Weighted Data.

|  |  | Week 7 | | | | Week 13 | | | | Week 52 | | | |
| --- | --- | --- | --- | --- | --- | --- | --- | --- | --- | --- | --- | --- | --- |
|  |  | TAU | RT | Diff. | *p** | TAU | RT | Diff. | p* | TAU | RT | Diff | p* |
| Raw data  M(SD) | ΔPCL-S | n=50 | n=226 |  |  | n=48 | n=211 |  |  | n=41 | n=191 |  |  |
|  |  | 20.22  (2.27) | 24.23  (1.27) | 4.01  (2.25) | 0.086 | 22.65  (2.43) | 26.16  (0.09) | 3.51  (2.40) | 0.220 | 23.21  (2.47) | 27.66  (1.36) | 4.45  (2.46) | 0.067 |
|  | ΔHSCL-25 | n=46 | n=213 |  |  | n=44 | n=200 |  |  | n=38 | n=177 |  |  |
|  |  | 0.41 (0.06) | 0.53  (0.03) | 0.12  (0.07) | 0.149 | 0.55 (0.07) | 0.60  (0.03) | 0.05 (0.08) | 0.561 | 0.56 (0.07) | 0.68  (0.03) | 0.12 (0.08) | 0.281 |
| Imputed,weighted data  M(SD) |  | n=70 | n=262 |  |  | n=70 | n=262 |  |  | n=70 | n=262 |  |  |
|  | ΔPCL-S | 28.29 (2.05) | 33.40  (0.09) | 5.11 (2.50) | 0.041 | 31.18  (2.48) | 36.31 (0.10) | 5.13 (3.26) | 0.157 | 35.02 (1.68) | 38.14  (0.10) | 3.12 (2.55) | 0.297 |
|  | ΔHSCL-25 | 0.48 (0.07) | 0.53  (0.03) | 0.05 (0.07) | 0.481 | 0.47  (0.07) | 0.50  (0.08) | 0.03 (0.11) | 0.621 | 0.64 (0.06) | 0.69  (0.04) | 0.05 (0.07) | 0.615 |

**p*-value were obtained using regression models adjusted on group and on the baseline value of the PTSD Checklist (PCL-S) total score or the Hopkins Symptom Checklist (HSCL-25) total score and, for imputed data weighted on propensity score after multiple imputations.
